# Physiotherapist and Patient Perspectives on a Snack-based Physical Activity Application and Tracking Device for People with Chronic Non-specific Low Back Pain: A Qualitative Study

**DOI:** 10.64898/2026.04.29.26351862

**Authors:** Ali Alali, Andrew Soundy, Deborah Falla, Janet Deane

**Affiliations:** Centre of Precision Rehabilitation for Spinal Pain (CPR Spine), School of Sport, Exercise and Rehabilitation Sciences, College of Life and Environmental Sciences, University of Birmingham, Edgbaston, Birmingham, B15 2TT, United Kingdom

**Author notes:** **Corresponding author:** Ali Alali, School of Sport, Exercise and Rehabilitation Sciences, University of Birmingham, Edgbaston, Birmingham, B15 2TT, United Kingdom, **Email:****, **.

**Keywords:** Low Back Pain, Chronic Pain, Exercise, Physical Activity, Qualitative Research, Mobile Applications, Telemedicine, Patient Compliance, Physical Therapists, Snack-based Physical Activity

## Abstract

**Objectives:** To explore patients’ and physiotherapists’ perspectives on a snack-based physical activity (PA) approach and mobile health technologies (mHealth) for non-specific chronic low back pain (NSCLBP). Snack-based PA refers to short, frequent bouts of activity (2–5 minutes) integrated into daily routines.

**Design:** Qualitative study using Interpretative Phenomenological Analysis (IPA) of semi-structured online interviews.

**Setting:** Community-based recruitment in the United Kingdom. Interviews were conducted online via Microsoft Teams between May and November 2024.

**Participants:** Sixteen participants were purposively sampled: eight adults with NSCLBP (lasting ≥3 months in the previous year) and eight physiotherapists with ≥2 years’ experience managing people with NSCLBP.

**Results:** Three shared themes were identified across both groups: (1) understanding the needs and requirements of PA; (2) perceptions of snack-based activity; and (3) factors influencing mobile health application use. Five subthemes were identified within themes one and three, together with two additional subthemes reported only by patients, relating to data sharing and technical issues. Both groups valued the time-efficiency and practical integration of snack-based activity, while highlighting the need for personalisation, age-appropriate content, accessibility and affordability.

**Conclusions:** Physiotherapists and patients emphasised the potential value of the snack-based PA approach in terms of adherence. However, both groups agreed that future intervention development should prioritise personalisation, user-friendly design, and equitable digital access.

**STRENGTHS AND LIMITATIONS OF THIS STUDY:**

- This study uses Interpretative Phenomenological Analysis (IPA) to provide in-depth, dual-perspective insights from both people with non-specific chronic low back pain (NSCLBP) and physiotherapists on a novel snack-based physical activity approach for chronic non-specific low back pain.
- Reporting adhered to the Consolidated Criteria for Reporting Qualitative Research (COREQ) 32-item checklist to ensure methodological transparency.
- The sample included variation in patient age, ethnicity and physiotherapist experience, which broadened the range of views captured.
- All participants were UK-based, which may limit the transferability of findings to other healthcare systems and cultural contexts.
- The small sample size, although consistent with IPA methodology, and the use of single online interviews, may not capture longitudinal perspectives on sustained engagement.

## INTRODUCTION

Non-specific low back pain (NSLBP) is defined as low back pain not attributable to a known cause [1]. Chronic NSLBP (NSCLBP) refers to any low back pain that persists beyond 12 weeks [2]. NSCLBP commonly hinders patients’ mobility, reduces physical activity [3] and is the leading cause of years lived with disability worldwide, profoundly impacting the quality of life [4]. In the UK, low back pain accounts for £12 billion in annual costs when healthcare and productivity losses are combined [5]. NSCLBP management remains a challenging issue due to its complex and multifactorial nature [6–8]. A holistic and multidisciplinary approach is required for the management of NSCLBP due to the interactions among various factors such as sedentary lifestyle and reduced physical activity levels, psychological features, and social factors [2, 9]. The National Institute for Health and Care Excellence (NICE) guidelines (NG59) endorse exercises in all forms such as stretching, strengthening or aerobic exercise like walking as a first-line treatment for chronic LBP, highlighting its efficacy in reducing pain and enhancing functional outcomes [10]. However, there is no clear guidance on the specific exercises that people should undertake to manage their NSCLBP since systematic reviews comparing various types of exercise programs have concluded that no exercise is better than any other for relieving chronic LBP [10, 11].

PA is defined as any ‘bodily movement produced by skeletal muscles that results in energy expenditure’ and the UK Chief Medical Officers’ guidelines (CMO) for the public recommend that adults should undertake at least 150 min of moderate to vigorous intensity of PA per week, supplemented by muscle-strengthening exercises twice weekly [12]. Notably, the 2019 update to the CMO guidance emphasises that every movement counts – adults no longer need to do exercise in bouts of 10 minutes or more for it to count toward the total, as even short bouts of activity confer health benefits [12]. Despite these recommendations, adherence to regular exercise remains low for people with NSCLBP. Some studies suggest that up to 70% of individuals with NSCLBP do not consistently follow their recommended home exercise regimens [13, 14]. Common reasons for low adherence include discomfort, low motivation, and time constraints [14]. In contrast, people with NSCLBP are more likely to adhere when they perceive a sense of mastery, ease, and perceived benefit [15]. Therefore, the management of people with NSCLBP via exercise needs to consider the constraints and limitations of traditional exercise programs and consider an innovative and creative approach.

One emerging intervention that could address exercise adherence in the NSCLBP population is a snack-based activity approach. Snacktivity™ is a novel approach that encourages incorporating small, brief and frequent activity bouts of exercise (2-5 minutes each) within the daily routine in inactive adults and the general population, yet the approach has not been tested on patients with NSCLBP [16]. Results have already shown that participation in snack-based PA results in increased PA and reduced sedentary episodes for the general population and inactive people [17–20]. The Snacktivity^TM^ approach includes physical activities such as climbing stairs, walking while meeting with colleagues, squatting while brushing your teeth or going for a brisk walk, which could be more easily integrated throughout the day to enhance PA level. This approach addresses common barriers, such as time constraints, and can make PA more approachable and accessible to sedentary people. However, whether this approach could be applied to people with NSCLBP remains unknown.

There is evidence to promote PA. The use of wearable smart technologies, such as tracking devices or telemedicine platforms, provides benefits to both patients and healthcare providers. Feedback from wearable smart technologies have demonstrated to increase PA levels by 20% in populations with chronic disease [21], as well as for chronic pain populations, for instance, a systematic review by Pfeiffer et al, [22], on the effectiveness of mobile health apps, reviewed 22 studies including 4679 participants and showed a small effect size (d = −0.40) in support of mobile health apps and their effect on chronic pain management in comparison to baseline measures or control interventions. These impacts were significant in the long term and more beneficial for chronic or recurrent pain patients [22].

Additionally, recent NICE guidance (2024) evaluated digital health technologies (e.g. getUBetter, SelfBack), telemedicine platforms and other mobile apps to improve outcomes for people suffering from NSCLBP, highlighting their potential to improve outcomes [23]. The use of these tools alongside standard treatment was shown to reduce pain, improve physical function, and increase patient autonomy by supporting self-management. Economic analysis showed cost-effectiveness and potential for healthcare resource savings, primarily through physiotherapy referrals, GP appointments, and reduced drug use. However, adherence issues and digital accessibility remain barriers and require careful consideration for equitable implementation [23].

In order to determine the content and usability of physical activity and Snacktivity™ qualitative research is required. Currently, there is a lack of qualitative information regarding the views and perceptions of this. Survey data has demonstrated the value of Snacktivity™ as a concept and the ability to self-monitor is important [18]. Review evidence has identified support for this and identification that Snacktivity™ is feasible, useable and acceptable across population groups [24]. However, to the best of the authors knowledge no past evidence has considered the perceptions of patients with chronic lower back pain.

This study aims to address critical gaps in adherence, accessibility and usability by exploring the perspectives of people with NSCLBP and physiotherapists perception of snack-based physical activity approach and the use of mobile health technologies in potentially improving outcomes among people with NSCLBP. The objective is to capture these perspectives, using semi-structured online interviews analysed by an interpretive phenomenological approach (IPA) [25]. This knowledge may help develop a tailored Snacktivity™ mobile health technology to enhance adherence and accessibility for patients with NSCLBP.

## METHODS

### Study design

The methodology employed in this study is based on the interpretive phenomenological analysis (IPA) [26]. This approach is helpful in investigating the experiences of the participants, making it possible to explore the nature of their pain, which is particularly relevant for persistent pain and complex bio-psychosocial connections [27]. IPA is recommended for analysing the lived reality of chronic pain as it is more flexible and responsive when compared with other theory-driven approaches [27]. Moreover, IPA employs hermeneutics, or interpretative theory, to understand how researchers interpret participants’ experiences [28]. In addition, IPA facilitates the exploration of the complexities of clinical decision-making, which requires advanced knowledge and awareness of a topic [29]. This qualitative study is reported in accordance with the Consolidated Criteria for Reporting Qualitative Research (COREQ) [30].

### Reflexivity

The first author (AA, BSc, MSc, PhD candidate) is a male physiotherapist experienced in NSCLBP management. AA completed IPA methodology training under Dr Soundy at the University of Birmingham, which enhanced the interpretation of participants’ responses to the semi-structured interview. AA had no prior relationship with any of the participants. To enhance reflexivity, field notes were made during and after each interview, then reviewed with co-authors to explore further interpretation and minimise bias.

### Participants

Inclusion criteria for people with NSCLBP:

- Males or females ≥ 18 years of age with experience of NSCLBP which lasted ≥ three months within the past year
- Capable of answering and understanding interview questions asked in English.

Inclusion criteria for the physiotherapists:

- Male or female qualified physiotherapists.
- Experience ≥ 2 years in treating people with NSCLBP
- Capable of answering and understanding interview questions asked in English.

### Sampling and sample size

In accordance with IPA studies, a small number of participants and homogeneous samples (typically less than 10) enabled a detailed and reflective analysis of the participants’ experience [26]. For this reason, two groups, including people with NSCLBP and physiotherapists with experience treating NSCLBP, were purposively sampled using a maximum variation approach to capture rich and in-depth accounts of individual lived experience, rather than thematic saturation, consistent with IPA guidance.

### Patient and public involvement

Prior to the design of this study, an informal patient and public involvement and engagement (PPIE) discussion was held in person at the University of Birmingham with approximately seven people living with non-specific chronic low back pain (NSCLBP) from the Centre of Precision Rehabilitation for Spinal Pain (CPR Spines) patient and public involvement community. The meeting was exploratory rather than structured, and sought patients’ views on the overall PhD project and specifically on the acceptability of a snack-based physical activity approach for NSCLBP. Their perspectives informed the study’s focus on acceptability and perceived value of the intervention and contributed to the framing of the interview topic guide. The interview topic guide was subsequently piloted with two individuals with NSCLBP and two physiotherapists experienced in NSCLBP management to ensure face validity; no changes were made as a result of the pilot. Patients and members of the public were not involved in participant recruitment or the conduct of the study. Results will be disseminated to study participants through a lay summary available from the corresponding author on request, and to the wider public through peer-reviewed publication and conference presentations.

### Data collection

Ethical approval was granted by the School of Science, Technology, Engineering and Mathematics Committee at the University of Birmingham (ERN_1890-Apr2024). Participants were recruited from the University of Birmingham staff and student population and from the wider Birmingham community (e.g. private practices). Recruitment methods included information posters displayed across University of Birmingham departments, email invitations, social media (Facebook, Instagram and X) and word of mouth. Interested participants were invited to undertake one online semi-structured interview based on open-ended questions. In advance of the interview, participants were asked to read an information sheet, which introduced the researcher and outlined the aims of the study, and to provide consent.

All interviews were conducted virtually on Microsoft Teams software hosted by the University of Birmingham between May and November 2024. The interviews lasted approximately 60 minutes and were conducted by a single researcher (AA). Demographic data were documented from each group including age, sex and ethnicity and for those with NSCLBP, this was supplemented by self-report questionnaires including their back pain intensity using a Numerical Pain Rating Scale (NRS) (0= no pain; 10= worst imaginable pain) [31], the Oswestry Disability Index (ODI) ( 0–100, higher scores indicating greater disability) and an International Physical Activity Questionnaire (IPAQ) to measure their current physical activity status (low, moderate, high physical activity level) [32, 33]. To confirm face validity, the interview schedule was piloted on two individuals with NSCLBP and two physiotherapists with expertise in CLBP management, no changes were made to the interview schedule following piloting. All online interviews were audio-recorded and transcribed verbatim using Microsoft Teams software to ensure accuracy and transparency. All interview transcripts were analysed manually using the IPA framework. Each participant was interviewed once, with no other individuals present, and no repeat interviews were conducted. Transcripts were not returned to participants for comment, and participants did not provide feedback on the findings.

### Data analysis

Each transcript was analysed separately to explore the uniqueness of each case and experience before making more general interpretations. The transcripts were read and analysed several times by the lead author (AA) only, using the IPA framework [26]. As the lead author read the transcript, initial notes were recorded in the margin of each transcript as required. Emerging themes were developed for each transcript by grouping the initial notes. Following the development of themes, the same researcher reviewed them to establish connections among the emerging themes. The emerging themes from the previous case were bracketed as the researcher analysed each participant’s transcript. Following the analysis of different case transcripts, the researcher conducted a cross-case analysis and began clustering the shared themes based on similarities in concepts, giving the group category descriptions relevant to the purpose and aim of this study. The last step was to interpret and review all of the themes systematically, searching for patterns and connections, allowing new superordinate themes to emerge.

To ensure quality in the approach, we adhered to established criteria, including constructing a coherent narrative, developing a vigorous phenomenological account, conducting close analytic readings of participants’ words, and addressing both convergence and divergence [34].

## RESULTS

### Demographic Characteristics and Clinical Outcome Measures

A total of 16 participants were involved in the study. Eight participants with NSCLBP completed the interview, as one participant withdrew prior to completion due to personal time constraints. The mean age of the participants with NSCLBP was 56.5 years (SD = 11.9). The sample consisted of five females (62.5%) and three males (37.5%), from a heterogeneous ethnic background. Participants reported experiencing NSCLBP for a mean duration of 10.7 years (SD = 9.5). Detailed participant demographics are presented in Table 1. The patients’ baseline clinical characteristics, including pain intensity measured by the NRS, PA levels assessed by the IPAQ, and disability related to LBP evaluated using the ODI, are summarised in Table 2.

**Table 1.** Participants’ (patients) demographics of age, gender, ethnicity and LBP history.

| Participants Demographics-Patients |  |  |  |  |
| --- | --- | --- | --- | --- |
| Participant ID | Gender | Age range (years) | Ethnicity | LBP History (years) |
| NSCLBP_1 | Female | 61–65 | White British | 30 |
| NSCLBP_2 | Female | 66–70 | White British | 2 |
| NSCLBP_3 | Male | 46–50 | Caucasian | 4 |
| Participant ID | Gender | Age range (years) | Ethnicity | LBP History (years) |
| NSCLBP_4 | Female | 56–60 | Black British | 16 |
| NSCLBP_5 | Female | 46–50 | Latino | 10 |
| NSCLBP_6 | Male | 46–50 | Chinese | 3 |
| NSCLBP_7 | Female | 76–80 | White British | 17 |
| NSCLBP_8 | N/A | N/A | N/A | N/A |
| NSCLBP_9 | Male | 36–40 | Black African | 3 |

**Table 2.** Participants’ (patients) characteristics and outcome measures of ODI, IPAQ and NRS.

(Table 1) Participants' (patients) demographics of age, gender, ethnicity and LBP history.
| Participants Characteristics-Patients |  |  |  |
| --- | --- | --- | --- |
| Participant ID | ODI (%) | IPAQ Category | NRS score (0 – 10) |
| NSCLBP_1 | 12 | High | 7/10 |
| NSCLBP_2 | 16 | Moderate | 4/10 |
| NSCLBP_3 | 24 | Low | 3/10 |
| NSCLBP_4 | 72 | Moderate | 9/10 |
| NSCLBP_5 | 38 | Low | 8/10 |
| NSCLBP_6 | 16 | High | 2/10 |
| Participant ID | ODI (%) | IPAQ Category | NRS score (0 – 10) |
| NSCLBP_7 | 36 | Low | 5/10 |
| NSCLBP_8 | N/A | N/A | N/A |
| NSCLBP_9 | 36 | Low | 2/10 |

Eight physiotherapists participated in the study. The physiotherapists had a mean clinical experience of 5.6 years (SD = 3.4), and possessed varied levels of qualifications, including bachelor’s, master’s, and doctoral degrees, ensuring heterogeneity within the sample. Further details about the physiotherapists’ demographics and qualifications are presented in Table 3.

**Table 3.** Participants’ (Physiotherapists) professional characteristics, such as years of experience and qualifications.

(Table 2) Participants' (patients) characteristics and outcome measures of ODI, IPAQ and NRS.
| Professional Characteristics of Physiotherapists |  |  |  |
| --- | --- | --- | --- |
| Participant ID | Chartered Physiotherapist | Experience (Years) | Qualification |
| PT_1 | Yes | 5 | MSc PT |
| PT_2 | Yes | 2 | Bachelor PT |
| PT_3 | Yes | 8 | MSc PT |
| PT_4 | Yes | 3 | MSc PT |
| PT_5 | Yes | 5 | MSc PT |
| PT_6 | Yes | 4 | Bachelor PT |
| PT_7 | Yes | 5 | Bachelor PT |
| PT_8 | Yes | 13 | PhD PT |

### Themes

A total of three main shared themes were identified: (1) Understanding the needs and requirements of PA, (2) Perception of snacks for PA and (3) Factors that influence the use of the mobile health app and recommendations. Themes and subthemes are summarised in Table 4. For clarity, participants with NSCLBP are labelled NSCLBP1–NSCLBP9 and physiotherapists are labelled PT1–PT8.

**Table 4.** Themes and subthemes are summarised in this table, with quotations from patients and physiotherapists.

| Themes and subthemes summary |  |  |  |
| --- | --- | --- | --- |
| Theme | Sub-theme | Group | Key Quotes |
| 1. Understanding the needs and requirements of physical activity | 1. Definition of PA | Both | **Patients:** "Anything that gets you up and moving basically." (NSCLBP_1)<br>**Physiotherapists:** "Any form of exercise that we can do during our days and just move our bodies for a specific amount of time." (PT_4) |
|  | 2. Low-moderate impact and activity incorporated into an individual's daily routine | Both | **Patients:** "Walking, pilates, and I do a healthy hearts aerobic class for an hour a week." (NSCLBP_2)<br>**Physiotherapists:** "Related to just activities of daily life, like taking stairs or going for a walk." (PT_2) |
|  | **3. Guidelines and personalised guidance for physical activity** | Both | **Patients:** "I think they need to consider the person as a whole rather than as someone on a piece of paper and one size fits all because it doesn't." (NSCLBP_4)<br>**Physiotherapists:** "The patient's preferences, expectations, and age; and the dose of exercise that is recommended." (PT_2) |
| **2. Perception of snacks for PA** | N/A | Both | **Patients:** "I think it's brilliant. Absolutely brilliant." (NSCLBP_4)<br>**Physiotherapists:** "It can break down patient sessions into smaller sessions, and I think this is really effective in managing my waiting list." (PT_8) |
| **3. Factors that influence the use of the mobile health app and recommendations** | **1. Ability to monitor progress and motivate** | Both | **Patients:** "I love statistics and data from exercise. I like to see what my heart rate is... and they have all these lovely graphs and statistics that show that which I really like." (NSCLBP_6)<br>**Physiotherapists:** "Because it motivates them... They can see their own progress." (PT_7) |
|  | **2. The need for age-specific content** | Both | **Patients:** "I think it should be specific for an age group because if you see somebody young and fit doing it, it's like, well, I can't do that." (NSCLBP_7)<br>**Physiotherapists:** "In the older population... using an app and you're telling the device to do something... I don't think it would be taken well." (PT_6) |
|  | **3. The need for accessibility, inclusivity, and affordability** | Both | **Patients:** "The price can be a big barrier as well." (NSCLBP_5)<br>**Physiotherapists:** "The price may pose a problem, especially if they<br>need to purchase a wearable device on top of the app." (PT_2) |
|  | **4. Data Sharing** | Patients Only | "I tend to stick to use things that have to do with Apple because I find far too many apps want your data and I'm not comfortable about that." (NSCLBP_4) |
|  | **5. Technical Issues** | Patients Only | "The only thing with it is it loses sync quite often, which is a bit annoying." (NSCLBP_1) |

#### Theme 1: Understanding the needs and requirements of physical activity

This theme identifies how patients and physiotherapists define PA, what types of PA are important and what considerations are needed around the promotion of PA and guidance.

#### Sub-theme 1: Definition of PA

Patients and physiotherapists shared a unified understanding of PA. They both perceived PA broadly as any type of movement. NSCLBP_1 stated that PA was *“anything that gets you up and moving basically,”* which demonstrated understanding of the term. Similarly, NSCLBP_3 highlighted that PA was *“anything from moving any part of my body, even stretching,”* demonstrating how even minimal activity can be considered as PA by patients. PT_4 described it as *“any form of exercise that we can do during our days and just move our bodies for a specific amount of time.”*. PT_8 shared a similar opinion stating, *“For me, physical activity means any movement.”* This shared understanding implies a potential common ground for patient-physiotherapist conversations about physical activity.

#### Sub-theme 2: Low impact and activity incorporated into an individual’s daily routine

Both physiotherapists and patients highlighted the value and importance of activities of daily living as an indicator of PA (physiotherapists) or as a preferred format of PA (patients). Patients showed a preference for low-impact activities such as walking, Pilates, and yoga, which were deemed helpful. NSCLBP_2 noted, *“Walking, Pilates, and I do a healthy hearts aerobic class for an hour a week,”* while NSCLBP_1 found stretching particularly useful, stating, *“Doing the stretching in yoga is really helpful,”* and highlighted the significance of walking: *“If I didn’t walk, I think I’d be in real trouble.”* Physiotherapists suggested integrating PA into the routine of daily living as an efficient strategy. PT_2 noted that physical activity can be *“related to just activities of daily life, like taking stairs or going for a walk.”*, PT_5 recommended *“walking and housework, things like that,”* suggesting that everyday activities can be effective in NSCLBP management. PT_8 also supported this, stating *“When I prescribe walking, they always come back to me and say they’re feeling better, they’re feeling good”*. There was an agreement on the preference for walking from both participant groups, since it is accessible and has perceived benefits.

#### Sub-theme 3: Guidelines and personalized guidance for physical activity

Both physiotherapists and patients referred to the guidelines and guidance of PA. However, this sub-theme demonstrated a distinction between the groups with physiotherapists focusing on guidelines and patients identifying the importance of personal needs, lifestyle and limitations to be accounted for. Patients suggested that a need for personalised PA was important to consider, and the needs should be based on preference and personal considerations like limitations caused by pain. NSCLBP_3 highlighted several considerations for physiotherapists stating *that “the lifestyle of each person because some activities are not doable or are not likeable for certain people or not possible for certain people.”* NSCLBP_4 shared a similar opinion, *“I think they need to consider the person as a whole rather than as someone on a piece of paper and one size fits all because it doesn’t.”* Pain was a significant limiting factor. Patients reported that their NSCLBP had decreased their ability to participate in PA. NSCLBP_2 stated, *“Yes, it’s reduced the intensity of it,”* when asked if back pain had affected the ability to undertake PA. NSCLBP_6 expressed caution due to pain, saying, *“I have a lower back pain that often happens if I lift a weight from the floor… Because I’ve got this low back pain, I’m very careful.”* Physiotherapists focused on identifying guidelines as evidence to guide their practice. For instance, PT_8 mentioned using *“ACSM (American College of Sports Medicine) guidelines, which state that there is a minimum requirement for each individual for physical activity criteria for each week.”* PT_6 was aware of the “WHO (define this here in brackets) guidelines for physical activity,” and PT_2 recalled guidelines suggesting *“a minimum of 150 minutes of physical activity per week.”* These statements indicate that for some physiotherapists, reference to guidance is important in terms of PA prescription. However, for the majority there was no reference to guidance to support their practice. Physiotherapists also emphasised that PA prescriptions should be individualised based on the patient requirement and preference. PT_2 emphasised the importance of factors such as *“the patient’s preferences, expectations, and age; and the dose of exercise that is recommended.”* PT_7 said they could assess factors like *“the weight of the person, looking [at] the gender… and consider their nutrition.”* Such individualisation of NSCLBP patient care could result in better patient outcomes and adherence.

#### Theme 2: Perception of snacks for PA

Both patients and physiotherapists agreed on the importance of the element of time efficiency by using a snack-based approach. For physiotherapists this could have implications for the NHS, waiting lists and better outcomes and for patients it had benefit around being able to incorporate PA into daily life. Patients found that engaging in daily PA snacks was appropriate. NSCLBP_4 appeared enthusiastic about the concept, expressing that, *“I think it’s brilliant. Absolutely brilliant.”* NSCLBP_6 shared a similar response, indicated that it could make it easier to integrate PA as part of a daily routine: *“I think it’s really good. If there’s anything that is a fix that doesn’t take a lot of time for someone who’s working full time, that’s a good idea.”* Patients valued the potential that PA snacks may offer in terms of not requiring additional time or impractical commitment. NSCLBP_6 highlighted this by saying, *“I like the fact that it’s short, that it’s instructional, so you don’t have to think about it… I like the idea that it’s not dependent on you having lots of time.”* NSCLBP_9, who works from home, found it practical to *“stretch a little bit and walk”* after sitting for several hours. Physiotherapists identified that one key benefit of implementing PA snacks is reported as the potential to improve the efficiency of physiotherapy services. PT_3 highlighted, *“If we prescribe physical activity, in that case they can continue for a longer time without our supervision. That’s why automatically the frequency to go to the hospital is decreasing.”* PT_8 stated, *“I think it’s absolutely efficient in working physiotherapy service because I would make more slots for patients and I would need less time with patients….It can break down patient sessions into smaller sessions, and I think this is really effective in managing my waiting list,”* These statements indicate that PA snacks could be used to improve the allocation of resources within Physiotherapy services. Employing PA snacks was also perceived as a way reduce the waiting list in physiotherapy clinics, making more time for other patients. PT_4 stated, *“This way most of the people might start seeing some improvement and this can reduce the number of patients that need to wait to be seen by a professional in the NHS.”* PT_8 added, *“It can break down patient sessions into smaller sessions and I think this is really effective in managing my waiting list.”* This implies that PA snacks could allow physiotherapists to manage their time more effectively by giving patients the tools to engage in self-directed activity.

#### Theme 3: Factors that influence the use of the mobile health app and recommendations

This theme identifies 5 sub-themes identified as factors likely to have a positive impact on patients. Sub-themes 1, 2, and 3 were common amongst both physiotherapists and patients. The last two were mentioned specifically by patients.

#### Sub-theme 1: Ability to monitor progress and motivate

The ability to monitor progress during activity and afterwards visualise the progress was highlighted by patients as valuable. Physiotherapists also identified the value of monitoring activities and associated this with goal setting. Patients and physiotherapists acknowledged the usefulness of mobile health apps for monitoring progress, providing motivation and setting goals. NSCLBP_6 stated, *“I love statistics and data from exercise. I like to see what my heart rate is… and they have all these lovely graphs and statistics that show that which I really like”*, This suggests that data presentation may enhance patient engagement and adherence and could be useful for patient monitoring. PT_4 remarked, *“I really like that you can record any type of physical activity… it provides data about almost anything”*. Consideration of reward systems and visual data could be used to enhance motivation among NSCLBP patients. NSCLBP_7 found the rewards motivating and mentioned, *“I’d make sure I did the number of steps a day, had to make sure my heartbeat was going okay, and I had to burn off a certain number of calories.”* The use of visual feedback and rewards appears to encourage continued engagement with physical activity. Physiotherapists acknowledged the value of apps in terms of monitoring activities and setting goals. PT_5 appreciated features that *“collect my workout data every day,”* and PT_2 also found interest in tracking specific features *“seeing information about performance levels and being able to track the volume of work done during the week”* PT_4 suggested that it could be useful if the app could send *“motivational messages and… reminders”* and *“a lot of graphs to show to the person that they are actually improving.”* Therefore, it seems that such functionalities could be used to improve patient engagement with an app and facilitate physiotherapist insight. Motivational features in apps could be beneficial for CLBP patients. PT_7 said the apps brought an element of fun: *“Because it motivates them… They can see their own progress.”*. PT_6 said,*“ it gives you medals when you complete a particular goal that you have set,”* which could motivate patient-app engagement. PT_1 remarked, *“They were kind of like give you a trophy kind of thing. Well, not a real trophy, but kind of like if you have accomplished like 5 days of running in the week you have two days off”*. This suggests that motivational features could be used to improve patient interest and PA intervention adherence.

#### Sub-theme 2: The need for age-specific content

Patients acknowledged that PA apps should contain age-specific content, indicating that the app content would need to be tailored to the individual. For example, NSCLBP_5 suggested, *“When you enter the app and enter your age, there should be a program for 20-year-olds and a program for 80-year-olds.”* NSCLBP_7 agreed, stating, *“I think it should be specific for an age group because if you see somebody young and fit doing it, it’s like, well, I can’t do that.”* This may mean that users may find it more appealing and useful if the material in the app is age-specific. On the other hand, age was frequently cited as a barrier by physiotherapists, with older patients being reportedly less comfortable with technology. PT_1 remarked: *“People are at least middle age or even… older age… they don’t know how to install, let alone how to use the application on their own.”* PT_6 added, *“In the older population… using an app and you’re telling the device to do something… I don’t think it would be taken well.”* These comments support the fact that the PA app will need to be used in conjunction with a healthcare professional who can provide appropriate training and personalise the interventions to ensure that they receive appropriate training and that it can be adapted according to patient preferences.

#### Sub-theme 3: The need for accessibility, inclusivity and affordability

Patients emphasised the need for enhanced app accessibility and inclusivity. Features such as language options, visual aids, and representation of diverse ethnicities were alluded to. NSCLBP_1 proposed making the app *“more pictorial and maybe include a talking feature to translate into other languages.”* NSCLBP_4 expressed a desire for representation, saying, *“I would want to see a mixture of ethnicities within the app.”* Such inclusivity could help people with NSCLBP to feel more connected to the app and increase its usability across different demographics. Patients also shared a concern with regard to the cost of application use. *“The price can be a big barrier as well”* said NSCLBP_5 and NSCLBP_6 said *“Cost. If it costs money or if it’s a subscription, that can be quite expensive based on what you’ve already signed up for. So that would be the main one.”.* Physiotherapists shared the same concerns with regard to the financial implications for patients. Regarding the PA app and the tracking device, PT_2 said *“The price may pose a problem, especially if they need to purchase a wearable device on top of the app,”.* PT_5 suggested that apps *“should be free to use”*. Therefore, when utilising a PA app or the tracking device as an adjunct to physiotherapy care, it will be essential to be mindful of cost. Otherwise, the financial affordability of a PA app or the tracking device may act as a barrier to patient accessibility.

#### Sub-theme 4: Data Sharing

Patients highlighted some privacy concerns. NSCLBP_4 expressed discomfort, stating, *“I tend to stick to using things that have to do with Apple because I find far too many apps want your data and I’m not comfortable about that.”* NSCLBP_9 admitted, *“I’m a private person and I’m paranoid about my privacy with these apps.”* These issues point to the need to ensure that any application used for the purposes of patient management will need to ensure that personal data is protected.

#### Sub-theme 5: Technical Issues

Technical challenges of using tracking devices were also considered as barriers. NSCLBP_1 noted, *“The only thing with it is it loses sync quite often, which is a bit annoying.”* NSCLBP_6 mentioned discomfort, saying, *“so it sometimes doesn’t recognise it on my device and then I have to manually do that. So, it’s slightly annoying, but generally it’s pretty good” referring* to a tracking device. Therefore, ease of app use will be essential for successful adoption.

## DISCUSSION

This qualitative study explored patients’ and physiotherapists’ perspectives regarding the use of a snack-based PA mobile application. This study has provided specific insights into how patients with NSCLBP and physiotherapists perceive snack-based activity, understand physical activity needs, and the role of mobile health applications in enhancing exercise adherence and accessibility. The discussion will now consider the following three central areas for further consideration: 1) understanding the needs and requirements of PA, 2) perception of snacks for PA, and 3) factors that influence the use of the mobile health app and recommendations.

### Understanding the needs and requirements of physical activity

Both patients and physiotherapists showed a similar understanding regarding the definition of PA, however, they differed in how PA is operationalised in practice. This shared perception between both patients and physiotherapists regarding PA is important in improving patient outcomes in patients with chronic pain due to its support of clearer understanding and communication between the therapist and patient [15, 35]. PA is also recognised as a therapeutic intervention for patients with NSCLBP [11, 36]. However, physiotherapists in our study emphasised local and international guidelines as references to the management of NSCLBP, patients favoured a personalised approach towards engaging in PA, not a ‘one size fits all’ approach. This has been widely acknowledged by previous work, which demonstrated that patients prefer a personalised approach towards their medical condition [37]. Nevertheless, both groups in our study understood that walking is an essential and acceptable form of PA for the management of NSCLBP.

### Perception of snacks for PA

The second theme in this study emphasises the perceived benefits of snacks of PA. This theme highlights the feasibility of snacks as an option for PA. Patients and physiotherapists in our study agree on the time efficiency of snack-based activities. Snack-based activities offer a small and time-efficient habit, making it easier to implement compared to a large structured activity [38]. This is in agreement with earlier studies suggesting that snack-based activities were designed for time efficiency [39]. Indeed, research has shown success in implementing snack-based activities in various settings [17]. Also, it offers freedom in scheduling exercise rather than engaging in time-consuming traditional exercises. This is important since studies have shown that one of the factors of adherence to exercise is the ability to train in their own time at their own convenience [14]. Our findings extend previous work regarding Snacktivity™ by demonstrating the feasibility of snack as an option among people with NSCLBP.

### Factors that influence the use of the mobile health app and recommendations

The third theme concerns factors that influence the use of the mobile health app among patients and physiotherapists. These factors, such as app ability to monitor progress, age appropriateness, accessibility, and data sharing, have offered prospects towards the use of the mobile app. Mobile app use, when combined with remote follow-up, has previously shown increased patient adherence in patients with chronic pain [21, 40, 41]. This type of monitoring and feedback that the participants of our study have shown is perfectly in line with studies that have demonstrated behaviour change through reinforcement [42]. Patients have also identified some considerations regarding content and the visual design of the app, where content should be age-appropriate and reflect diverse users to enhance app inclusivity. These factors could affect usability, where some groups, such as older adults, may feel less confident using this technology. This is an important consideration given that there is an age ‘digital divide’ in healthcare [43]. Patients flagged privacy concerns regarding data sharing, which is considered a valid concern regarding the use of mobile health technologies, as stated elsewhere [44, 45]. Therefore, there is a need to protect data, and adopting secure technology is important when developing mobile health technologies.

### Methodological considerations

This study has several strengths, including two important groups associated with NSCLBP and the use of the IPA approach to obtain in-depth analyses of patients’ and physiotherapists’ perceptions regarding the snack-based PA application and the use of a tracking device. However, there are some limitations associated with this study. Firstly, the demographic homogeneity of both UK-based patients and physiotherapists included in the study limits the generalisability of the transferability of the findings to other regions and healthcare systems. Another limitation is that although the small sample size of the study is justified, given the depth of patient and physiotherapist responses required for this IPA analysis, it may still restrict the diversity of experiences captured; therefore, future studies could improve on this by adding a larger and more diverse sample. Whilst the focus of this current study was on the short-term effects of snack-based physical activities for NSCLBP within a small sample, future consideration of the long-term effects will be important for larger, longitudinal study designs across diverse patient and physiotherapist populations, countries and healthcare settings.

## Conclusion

Findings of this study have demonstrated the practicality and feasibility of a snack-based PA mobile app for patients with NSCLBP. Both patients and physiotherapists showed a similar understanding regarding the definition of PA, however, they diverged on how it could be operationalised in practice. Physiotherapists in our study emphasised local and international guidelines as references to the management of NSCLBP. Patients favoured a personalised approach towards engaging in PA, while both groups agreed on snack-based activity being time-efficient and an acceptable way of improving PA. However, there are slight concerns regarding patients’ privacy and app accessibility. Future research should address these concerns for better app development for implementation in the rehabilitation of people with NSCLBP.

## Supporting information

COREQ 32-item Reporting Checklist

Interview Topic Guide for Patients

Interview Topic Guide for Physiotherapists

## Data Availability

All data produced in the present study are available upon reasonable request to the authors.

## DECLARATIONS

## Ethics approval

This study was approved by the School of Science, Technology, Engineering and Mathematics Ethics Committee at the University of Birmingham (reference ERN_1890-Apr2024). All participants provided written informed consent.

## Contributorship statement

AA, AS, DF and JD conceived and designed the study. AA recruited participants, conducted the interviews and led the analysis. AS, DF and JD supervised the research and contributed to the interpretation. AA drafted the manuscript. All authors critically revised the manuscript, approved the final version, and agree to be accountable for all aspects of the work. AA is the guarantor.

## Data sharing statement

The de-identified data generated and analysed during this study are not publicly available to protect participant confidentiality. Anonymised extracts that support the findings may be made available from the corresponding author on reasonable request, subject to ethics committee approval.

## Acknowledgements

The authors thank all participants, both people with NSCLBP and physiotherapists, who generously gave their time to this study.

## Use of AI statement

The authors used ChatGPT (OpenAI) and Claude (Anthropic) to assist with language editing, formatting, and some structuring of administrative components of the manuscript. No identifiable participant data were entered into the tools. All outputs were critically reviewed and verified by the authors, who take full responsibility for the final manuscript.

## Patient consent for publication

Not applicable — no identifying patient information is reported. All quotations are de-identified.

## Provenance and peer review

Not commissioned, externally peer reviewed.

