## Supplementary material for "Physiotherapist and Patient Perspectives on a Snack-based Physical Activity Application and Tracking Device for People with Chronic Non-specific Low Back Pain: A Qualitative Study": COREQ 32-item Reporting Checklist

**COREQ (Consolidated criteria for Reporting Qualitative research) Checklist**

*A 32-item checklist for interviews and focus groups*

| **Topic** | **Item No.** | **Guide Questions / Description** | **Reported on Page No.** |
| --- | --- | --- | --- |
| **Domain 1: Research team and reflexivity** | | | |
| ***Personal characteristics*** | | | |
| Interviewer / facilitator | **1** | Which author/s conducted the interview or focus group? | p. 8 (Reflexivity); p. 10 (Data collection) — "...conducted by a single researcher (AA)." |
| Credentials | **2** | What were the researcher's credentials? E.g. PhD, MD | p. 8 (Reflexivity) — "AA, BSc, MSc, PhD candidate." |
| Occupation | **3** | What was their occupation at the time of the study? | p. 8 (Reflexivity) — "...male physiotherapist [and] PhD candidate." |
| Gender | **4** | Was the researcher male or female? | p. 8 (Reflexivity) — "male physiotherapist." |
| Experience and training | **5** | What experience or training did the researcher have? | p. 8 (Reflexivity) — "...experienced in NSCLBP management. AA completed IPA methodology training under Dr Soundy at the University of Birmingham..." |
| ***Relationship with participants*** | | | |
| Relationship established | **6** | Was a relationship established prior to study commencement? | p. 8 (Reflexivity) — "AA had no prior relationship with any of the participants." |
| Participant knowledge of the interviewer | **7** | What did the participants know about the researcher? e.g. personal goals, reasons for doing the research | p. 10 (Data collection) — "...participants were asked to read an information sheet, which introduced the researcher and outlined the aims of the study, and to provide consent." |
| Interviewer characteristics | **8** | What characteristics were reported about the interviewer/facilitator? e.g. bias, assumptions, reasons and interests in the research topic | p. 8 (Reflexivity) — "...field notes were made during and after each interview, then reviewed with co-authors to explore further interpretation and minimise bias." |
| **Domain 2: Study design** | | | |
| ***Theoretical framework*** | | | |
| Methodological orientation and theory | **9** | What methodological orientation was stated to underpin the study? e.g. grounded theory, discourse analysis, ethnography, phenomenology, content analysis | p. 7–8 (Methods – Study design) — "The methodology employed in this study is based on the interpretive phenomenological analysis (IPA)." |
| ***Participant selection*** | | | |
| Sampling | **10** | How were participants selected? e.g. purposive, convenience, consecutive, snowball | p. 9 (Sampling and sample size) — "...two groups...were purposively sampled using a maximum variation approach..." |
| Method of approach | **11** | How were participants approached? e.g. face-to-face, telephone, mail, email | p. 10 (Data collection) — "Recruitment methods included information posters displayed across University of Birmingham departments, email invitations, social media (Facebook, Instagram and X) and word of mouth." |
| Sample size | **12** | How many participants were in the study? | p. 12 (Results – Demographic Characteristics) — 16 participants (8 with NSCLBP; 8 physiotherapists). |
| Non-participation | **13** | How many people refused to participate or dropped out? Reasons? | p. 12 (Results) — "...one participant withdrew prior to completion due to personal time constraints." |
| ***Setting*** | | | |
| Setting of data collection | **14** | Where was the data collected? e.g. home, clinic, workplace | p. 10 (Data collection) — "All interviews were conducted virtually on Microsoft Teams software...between May and November 2024." |
| Presence of non-participants | **15** | Was anyone else present besides the participants and researchers? | p. 11 (Data collection) — "Each participant was interviewed once, with no other individuals present..." |
| Description of sample | **16** | What are the important characteristics of the sample? e.g. demographic data, date | p. 12–14 (Results, Tables 1–3) — age, sex, ethnicity, LBP duration, NRS, ODI, IPAQ for patients; clinical experience and qualifications for physiotherapists. Interview dates May–November 2024 (p. 10). |
| ***Data collection*** | | | |
| Interview guide | **17** | Were questions, prompts, guides provided by the authors? Was it pilot tested? | p. 10–11 (Data collection) — "To confirm face validity, the interview schedule was piloted on two individuals with NSCLBP and two physiotherapists with expertise in CLBP management; no changes were made to the interview schedule following piloting." |
| Repeat interviews | **18** | Were repeat interviews carried out? If yes, how many? | p. 11 (Data collection) — "Each participant was interviewed once...no repeat interviews were conducted." |
| Audio / visual recording | **19** | Did the research use audio or visual recording to collect the data? | p. 11 (Data collection) — "All online interviews were audio-recorded and transcribed verbatim using Microsoft Teams software..." |
| Field notes | **20** | Were field notes made during and/or after the interview or focus group? | p. 8 (Reflexivity) — "...field notes were made during and after each interview, then reviewed with co-authors..." |
| Duration | **21** | What was the duration of the interviews or focus group? | p. 10 (Data collection) — "The interviews lasted approximately 60 minutes..." |
| Data saturation | **22** | Was data saturation discussed? | p. 9 (Sampling and sample size) — "...purposively sampled using a maximum variation approach to capture rich and in-depth accounts of individual lived experience, rather than thematic saturation, consistent with IPA guidance." |
| Transcripts returned | **23** | Were transcripts returned to participants for comment and/or correction? | p. 11 (Data collection) — "Transcripts were not returned to participants for comment..." |
| **Domain 3: Analysis and findings** | | | |
| ***Data analysis*** | | | |
| Number of data coders | **24** | How many data coders coded the data? | p. 11 (Data analysis) — "The transcripts were read and analysed several times by the lead author (AA) only, using the IPA framework." |
| Description of the coding tree | **25** | Did authors provide a description of the coding tree? | p. 11 (Data analysis) — Stepwise IPA process: initial notes → emerging themes → cross-case clustering → superordinate themes. Final themes/subthemes presented in Table 4 (p. 22–24). |
| Derivation of themes | **26** | Were themes identified in advance or derived from the data? | p. 11 (Data analysis) — Themes were inductively derived from the data following the IPA framework. |
| Software | **27** | What software, if applicable, was used to manage the data? | p. 10–11 (Data collection & Data analysis) — Microsoft Teams used for recording/transcription. "All interview transcripts were analysed manually using the IPA framework." (No dedicated qualitative analysis software was used.) |
| Participant checking | **28** | Did participants provide feedback on the findings? | p. 11 (Data collection) — "...participants did not provide feedback on the findings." |
| ***Reporting*** | | | |
| Quotations presented | **29** | Were participant quotations presented to illustrate the themes / findings? Was each quotation identified? e.g. participant number | p. 15–22 (Results – Themes) — Verbatim quotations are presented throughout, identified by participant ID (NSCLBP_1–NSCLBP_9; PT_1–PT_8). |
| Data and findings consistent | **30** | Was there consistency between the data presented and the findings? | p. 15–22 (Results) and p. 24–26 (Discussion) — Themes and subthemes are supported by direct quotations and interpreted consistently in the Discussion. |
| Clarity of major themes | **31** | Were major themes clearly presented in the findings? | p. 15 (Results – Themes) — Three major themes clearly presented with subheadings: (1) understanding the needs and requirements of PA; (2) perception of snacks for PA; (3) factors influencing the use of the mobile health app and recommendations. |
| Clarity of minor themes | **32** | Is there a description of diverse cases or discussion of minor themes? | p. 15–22 (Results) — Five subthemes across themes 1 and 3, plus two patient-only subthemes (data sharing; technical issues) describing divergent views. |

*Developed from: Tong A, Sainsbury P, Craig J. Consolidated criteria for reporting qualitative research (COREQ): a 32-item checklist for interviews and focus groups. International Journal for Quality in Health Care. 2007. Volume 19, Number 6: pp. 349–357.*
