## Supplementary material for "Physiotherapist and Patient Perspectives on a Snack-based Physical Activity Application and Tracking Device for People with Chronic Non-specific Low Back Pain: A Qualitative Study": Interview Topic Guide for Patients

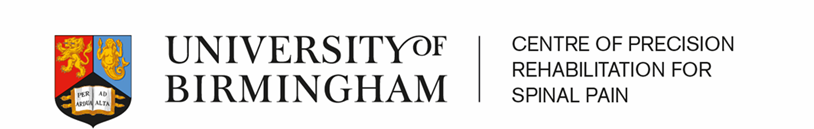

**Patient Interview Topic Guide**

**Study Title**:

Physiotherapist and Patient Perspectives on a Snack-based Physical Activity Application and Tracking Device for People with Chronic Non-specific Low Back Pain: A Qualitative Study.

**Aim of this interview:**

To evaluate patient perspectives on physical activity and mobile health technologies to improve outcomes for people with chronic non-specific low back pain.

We are grateful for your agreement to take part in this study and ask that you spend ten minutes thinking about the listed questions before the process begins.

This interview will take about 1 hour. I will be asking you some questions about physical activity and a physical activity app for people with non-specific chronic low back pain.

The interview questions aim to evaluate your views on physical activity and mobile health technologies to improve outcomes for people with chronic non-specific low back pain. There are no right or wrong answers, I am just interested in your honest opinions and perspectives.

You will retain your anonymity in this research and all identifiable information, such as your name, will not be recorded. I would like to audio record our discussion so I can focus on our conversation. The recording will only be accessed by the research team and deleted after the study. You can ask me to stop recording at any point. Thank you for taking part in this research. We will proceed when you are ready.

**Definitions**:

In this interview I will be mentioning ‘physical activity’, ‘nonspecific chronic low back pain’ and a ‘physical activity App or tracker’. I will now go through the definition of each but please feel free if you do not understand terms within this interview.

By physical activity I mean any activity that requires bodily movement (for example, daily activity, activities at work, housework, getting from place to place or active recreation and sport).

By ‘physical activity app’ or ‘tracker’ I mean mobile phone applications and physical activity tracking devices (i.e., fitbit) used to encourage and track physical activity.

By non-specific chronic low back pain, I mean persistent low back pain that has lasted for more than 3 months.

Section 1 Demographics

I will now ask you some questions in relation to your demographics to help us to understand how to develop an inclusive physical activity management approach for people with persistent low back pain. If you feel uncomfortable to answer any of the following questions, please let me know.

1. What age are you?
2. Can you confirm what your ethnicity is?
3. How long have your experienced persistent low back pain for?
4. Have you managed to answer the International Physical Activity Questionnaire? If not, are you happy to complete them at the end of the interview?

Section 2 Physical activity

1. What does physical activity mean to you?
2. Have you ever been prescribed physical activity to manage your low back pain?
3. Have you ever been advised to increase or maintain your general physical activity level as part of your low back pain treatment?
4. As someone who has experienced low back pain, is there a type of physical activity that you prefer or practice on a regular basis?
5. In your opinion, when physiotherapists are recommending or prescribing physical activity interventions, is there anything that they need to consider?
6. Would you describe yourself as a physically active person?
7. Has your low back pain reduced your physical activity?
8. Is there any particular type of physical activity that improves your low back pain?
9. Is there any particular type of physical activity that makes it worse?
10. What other factors, aside from pain, limits your physical activity?
11. What do you currently do for physical activity each week? How much activity do you do each week?
12. Do you think you do enough physical activity to stay healthy?

Section 3 Physical activity snacks

1. In this current study we are defining a ‘physical activity snacks’ for people with chronic low back pain as small bouts of daily physical activity that could be used to improve pain and disability for people with chronic pain. What do you think about this idea?
2. What do you like about it?
3. What don’t you like?
4. Would this approach help you to be more physically active? Why? Why not?
5. What would stop you doing physical activity snacks every day?
6. Given your experience of persistent low back pain, which physical activity snacks do you think you could fit into your daily life? [Prompt: Ask about work and home and provide examples of snack activity e.g., walk to work, walk upstairs, body movements].
7. As discussed, for the purposes of this study we are using the term ‘physical activity snack’ or ‘back snack’ to denote small bouts of physical activity for people with chronic low back pain. Do you like this term? If you don’t like it, why? Do you have any suggestions?

Section 4 Mobile health Apps

1. Do you use apps (applications on a mobile device or wearable). If yes, how often do you use them? If not, why not?
2. Which apps do you use?
3. Do you use apps to monitor or track your physical activity?
4. If yes, how do you find using your phone app to track and monitor your physical activity?
5. How easy was it to learn to use the app? How clear are the menu labels and instructions?
6. Is the app interesting to use? Does it use any strategies to keep you using the app?
7. How well does the app look?
8. Does it present information in an interesting way?
9. Would you recommend this app to others? If so, why? If not, why?
10. What features do you particularly like? Can you tell us why?
11. What features do you particularly dislike? Can you tell us why?
12. Has the app encouraged you to be more physically active? If so, please tell us why? If not- please tell us what you may find useful to encourage more physical activity.

**Section 5 Mobile apps for people with chronic low back pain**

1. Do you use apps to help manage your low back pain?
2. If so, which ones do you find useful? If not, why not?
3. What are the main barriers to app use? [Prompt: what inhibits you from using apps to help your low back pain]
4. What are the main facilitators? [Prompt: what encourages you to use an app to manage your low back pain]
5. If you were to design an app to help people with low back pain, what would it need to look like?
6. If you were to design an app to help people with low back pain, what would it contain?
7. If you were to design an app to help people with low back pain, how would you make it accessible for all? [Prompt: inclusive of all ethnic minority groups, age groups, genders].

Section 6 Mobile health tracking devices

1. Do you use tracking devices to monitor your activity?
2. What features do you like the most? What don’t you like about it?
3. Has your tracker helped you to be more physically active?
4. Has it helped you set activity goals? Has it helped you reach your activity goals more rapidly? Can you tell us why/ or why not?
5. Has it made it easier for you to be more active? In what ways?
6. Is it easy to learn how to operate the activity tracker?
7. Is it clear and understandable to use?
8. Is comfortable to wear?
9. Do you use a tracker device in association with the app? [Prompt: tracker or wearable device e.g., Fit bit]. If so, what do you think of it?
10. Many smartphone apps that track activity only work when you have your smartphone with you–do you always have your smart phone with you?

**Section 7 Further comments**

Do you have any further comments?
